# Highly versatile antibody binding assay for the detection of SARS-CoV-2 infection

**DOI:** 10.1101/2021.07.09.21260266

**Authors:** Pratik Datta, Rahul Ukey, Natalie Bruiners, William Honnen, Mary O. Carayannopoulos, Charles Reichman, Alok Choudhary, Alberta Onyuka, Deborah Handler, Valentina Guerrini, Pankaj K. Mishra, Hannah K. Dewald, Alfred Lardizabal, Leeba Lederer, Aliza L. Leiser, Sabiha Hussain, Sugeet K. Jagpal, Jared Radbel, Tanaya Bhowmick, Daniel B. Horton, Emily S. Barrett, Yingda L. Xie, Patricia Fitzgerald-Bocarsly, Stanley H. Weiss, Melissa Woortman, Heta Parmar, Jason Roy, Maria Gloria Dominguez-Bello, Martin J. Blaser, Jeffrey L. Carson, Reynold A. Panettieri, Steven K. Libutti, Henry F. Raymond, Abraham Pinter, Maria Laura Gennaro

## Abstract

Monitoring the burden and spread of infection with the new coronavirus SARS-CoV-2, whether within small communities or in large geographical settings, is of paramount importance for public health purposes. Serology, which detects the host antibody response to the infection, is the most appropriate tool for this task, since virus-derived markers are most reliably detected during the acute phase of infection. Here we show that our ELISA protocol, which is based on antibody binding to the Receptor Binding Domain (RBD) of the S1 subunit of the viral Spike protein expressed as a novel fusion protein, detects antibody responses to SARS-CoV-2 infection and COVID-19 vaccination.

We also show that our ELISA is accurate and versatile. It compares favorably with commercial assays widely used in clinical practice to determine exposure to SARS-CoV-2. Moreover, our protocol accommodates use of various blood- and non-blood-derived biospecimens, such as breast milk, as well as dried blood obtained with microsampling cartridges that are appropriate for remote collection. As a result, our RBD-based ELISA protocols are well suited for seroepidemiology and other large-scale studies requiring parsimonious sample collection outside of healthcare settings.

## INTRODUCTION

Diagnosis of infection with the novel coronavirus SARS-CoV-2, the causative agent of the ongoing COVID-19 pandemic, has relied on two classes of assays. One comprises the methods for detecting the presence of the virus in upper respiratory specimens, either by viral nucleic acid amplification tests (NAAT) or immunodetection of viral antigen. NAATs based on Real-time PCR represent the gold standard for diagnosis of acute SARS-CoV-2 infection while the antigen tests, which are comparatively less sensitive, are critically important for public health purposes, since they have a very rapid turn-around and detect infectious cases (1-6). The second class of assays comprises methods for detecting virus-specific antibodies in peripheral blood. These antibodies are reliable indicators of viral exposure, since they become detectable approximately two weeks after initiation of productive infection and typically persist for 6-12 months or longer, well beyond the time in which virus detection assays return to negativity (**Fig. 1**). Thus, antibody-based assays are most valuable as metrics of infection burden in the population for epidemiological purposes and large-scale studies.

**Figure 1.**
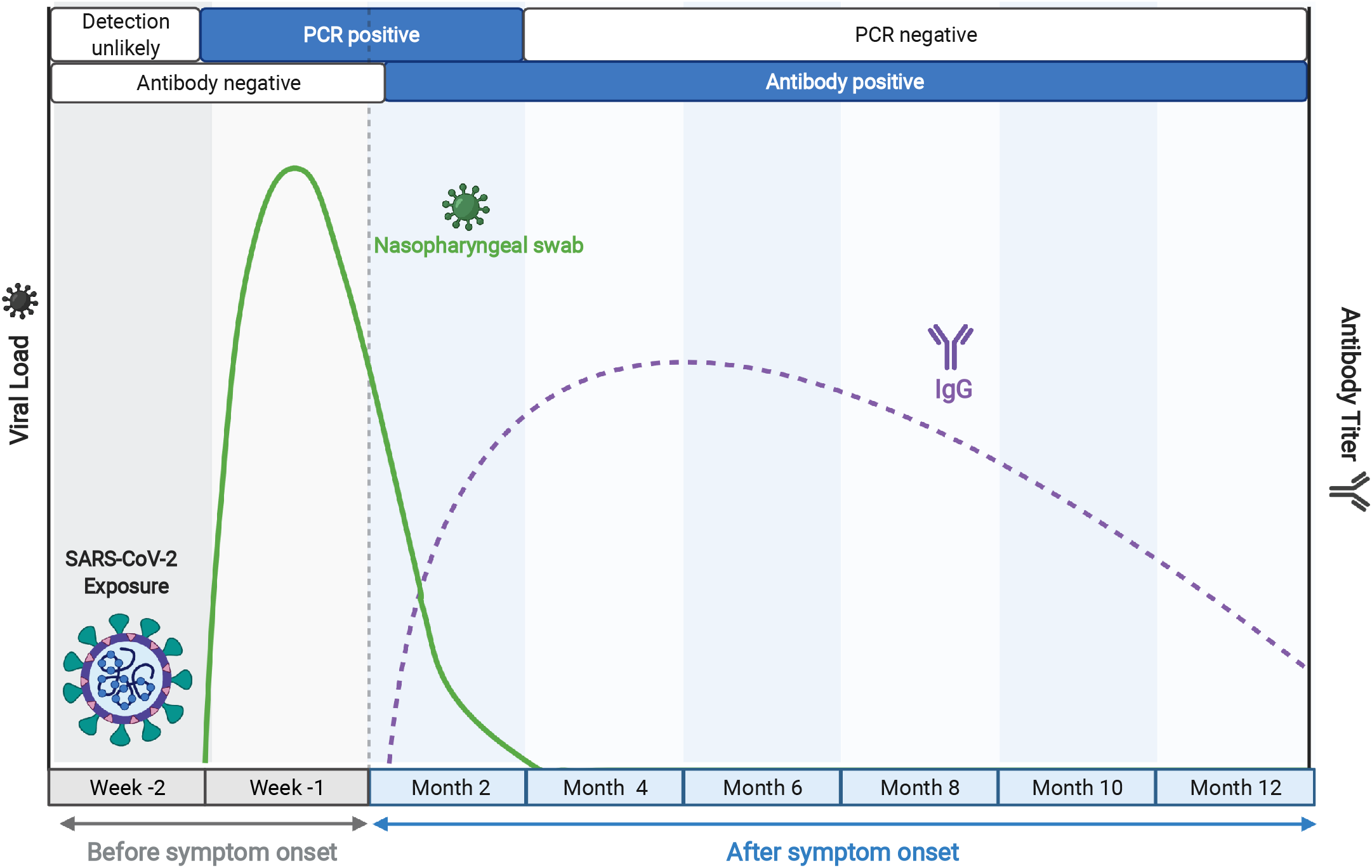
Time course of key biomarkers in SARS-CoV-2 infection, adapted from BioRender.com. The solid green line represents a typical trajectory of the RT-PCR data for viral nucleic acid from respiratory samples, while the broken purple line indicates a typical virus-specific antibody trajectory in peripheral blood, relative to time of infection, as indicated.

Antibody-based assays for SARS-CoV-2 infection are based on two SARS-CoV-2 antigens. One is Spike (S), a two-subunit protein that decorates the surface of the virion and establishes contact with the host cell receptor, angiotensin-converting enzyme 2 (ACE2), through the receptor-binding domain (RBD) in the S1 subunit, thus determining host range and tissue tropism (7). The second viral antigen is the Nucleocapsid (N), which interacts with the viral genomic RNA inside the viral envelope. Both antigens have been used for SARS-CoV-2 antibody detection, with a preference for the N antigen in most commercial antibody detection assays utilized in clinical settings (for example, (8, 9)). In contrast, the S protein has been mostly adopted as antibody capture antigen in research settings (e.g., (10-12)), primarily because the S1 RBD region is particularly immunogenic and the dominant target of neutralizing (protective) antibodies (11, 13, 14). Moreover, mutations in RBD, which are important factors in the evolution of all major SARS-CoV-2 variants, increase affinity for the ACE-2 receptor and lead to resistance to monoclonal and polyclonal antibodies developed in response to infection or vaccination (15-17). Furthermore, use of S1 RBD for antibody testing has been extended to clinical applications (see list of emergency use authorized serology tests at fda.gov/medical-devices) since the introduction of COVID-19 vaccines, which contain S but not N (18, 19). Thus, it becomes increasingly important to identify all potential uses of S-based serological assays for SARS-CoV-2 infection.

Here we describe key characteristics of our serological assay utilizing a novel S1 RBD antigen and its suitability for antibody detection from minimal (μl scale) amounts of remotely collected peripheral blood, which is critical for seroepidemiological and large-scale studies conducted outside of health care settings. We also show that the assay is equally suited for detecting antibodies in different liquid compartments of peripheral blood and other bodily fluids, making it adaptable to diverse study designs.

## MATERIALS AND METHODS

### Human Subjects Ethics Statement

The analyses presented in the present work draw upon data and biospecimens gathered during seven studies of COVID-19 in NJ, USA. All participants were enrolled after written informed consent was obtained from each participant. All study subjects were >20 years of age. To assess antibody responses to SARS-CoV-2 infection we used plasma/serum samples obtained from 83 SARS-CoV-2 PCR-confirmed convalescent subjects (20); 146 patients hospitalized for COVID-19 (PCR confirmed) at Robert Wood Johnson University Hospital in New Brunswick, NJ; blood collected after >2 weeks from completion of full vaccination from 283 subjects vaccinated against COVID-19 between mid-December 2020 and mid-February 2021 among healthcare workers in Rutgers-affiliated hospitals (21) and Rutgers employees (20); and 148 residents living in the township of Lakewood, NJ in April 2020. Studies were approved by the Research Subjects Institutional Review Board at the University of Rutgers, Newark, New Jersey (Pro2020000655, Pro2020001263, and ClinicalTrials.gov registration numbers NCT04336332 and NCT04336215). As negative controls, we used 104 stored serum/plasma samples collected prior to the COVID-19 pandemic (Institutional review board of the Rutgers New Jersey Medical School, Pro0119980237 and Pro20150001314) and 103 serum samples obtained during the pandemic from subjects who remained SARS-CoV-2 PCR-negative for at least 16 weeks following the blood draw utilized in the study (Pro2020000679 and ClinicalTrials.gov registration number NCT04336215). Breast milk was obtained from four SARS-CoV-2 PCR-negative lactating mothers (Pro2018002781), by hand expressing or pumping into sterile glass vials. All biospecimens were linked to de-identified study ID numbers.

### Expression and purification of recombinant SARS-COV-2 S1 RBD Protein

A DNA fragment encoding RBD (Spike residues aa. 316 to aa. 544) was amplified and cloned at the 3’ end of a gene expressing the N-terminal fragment of the Fr-MuLV SU (gp70 protein) in the eukaryotic expression vector pcDNA3.4 (Addgene, Watertown, MA). The resulting plasmid was transfected into 293F cells using the Expi293 Expression system (Thermo Fisher Scientific, Waltham, MA), according to the manufacturer’s protocol. Supernatants were collected on day 3 post-transfection, and recombinant protein was purified by absorption to HisPur™ Cobalt Resin (Thermo Fisher Scientific, Waltham, MA) and elution with 200mM imidazole. Purified protein was subsequently dialyzed against PBS at 4°C. Absorbance (OD_280_) was determined by Nanodrop reading and concentrations were calculated using the ExPASy Proteomics calculator. Molecular weights were adjusted to account for the number of N-linked glycosylation sites to determine the final concentration.

### Reference SARS-COV-2 S1 RBD Protein

The reference protein was produced from the vector pCAGGS containing the SARS-Related Coronavirus 2, Wuhan-Hu-1 Spike Glycoprotein Receptor Binding Domain (RBD) (Catalog No. NR-52309, BEI Resources, Manassas, VA), utilizing the manufacturer’s protocol.

### Non-SARS-CoV-2 antigens

Spike Protein S1 from non-SARS-CoV-2 coronaviruses (HCoV-229E, HCoV-NL63, HCoV-HKU-1) and Spike Protein S1 and S2 extracellular domain (HCoV-OC43) were obtained from Sino Biologicals (Wayne, PA, USA) and pooled in equimolar amounts to a final concentration of 1 mg/ml. The resulting pool was used at 2 µg/ml (50 μl per well) to coat the ELISA plates.

### Antibody binding by enzyme-linked immunosorbent assay (ELISA)

96-well ELISA plates (Nunc MaxiSorp, ThermoFisher, Rochester, NY) were coated with 2 µg/ml recombinant SARS-CoV-2 RBD (50 μl per well) overnight at 4°C. Plates were washed four times with 100 μl/well washing buffer (1x PBS containing 0.05% Tween 20) (Sigma-Aldrich, St. Louis, MO) and blocked with 100 μl/well blocking buffer [2% Blotto (Nestle Carnation, US) in PBS] for 30 min at 37°C. Diluted plasma/serum (1:1 in 1x PBS) was heat-inactivated at 56°C for 1 hour prior to use. After blocking, plates were washed four times with 100 μl /well washing buffer, and 50 μl plasma/serum diluted in blocking buffer was added to each well and incubated for 1 hour at 37°C. For matrix equivalence studies, serum was diluted in the test matrices (breast milk or plasma obtained from blood collected in various anticoagulant tubes), as described in *Results*. Bound IgG was detected by adding alkaline phosphatase-conjugated goat anti-human IgG (Jackson ImmunoResearch, West Grove, PA) diluted 1:2,000 in blocking buffer (50 μl/well). Enzyme activity was assayed by adding 50 μl/well phosphate substrate (Sigma-Aldrich, St. Louis, MO) solubilized in developing buffer (2:1 diethanolamine:MgCl2.6H_2_O (Sigma-Aldrich), pH 9.8). The reaction was stopped after 30 min with 1M NaOH (50 μl/well) and results were read as absorbance (OD_405_) values.

Each ELISA plate contained positive and negative serum/plasma controls and background control wells without primary antibody, and each sample was tested in duplicate. The protocol was automated, using a Hamilton Microlab STAR liquid handler (Hamilton Company, Reno, NV) for sample handling and dilution, and a BioTek EL406 combination washer dispenser and a Synergy Neo2 microplate reader (BioTek, Winooski, VT) for ELISA. Work involving blood products from SARS-CoV-2-infected subjects was performed in a biosafety level 2+ (BSL-2+) laboratory utilizing protocols approved by the Rutgers Institutional Biosafety Committee.

### Commercial antibody detection assays

The Roche Elecsys^®^ Anti-SARS-CoV-2 assay utilizing the Roche Cobas e601 instrument and the Abbott Architect SARS-CoV-2 IgG assay utilizing the Abbott Architect c4000, which both use SARS-CoV-2 N protein as capture antigen, were performed by specialized personnel following the manufacturer’s instructions.

### Sample Collection and Processing

For phlebotomy, standard venipuncture was performed, and 10 mL of blood was collected in a serum separator tube with inert clot activator (BD367861, Franklin Lakes, NJ) or in a tube containing the anticoagulant sodium heparin (BD366480). For the matrix equivalence study, tubes containing other anticoagulants [potassium/EDTA (BD367861), lithium heparin (BD367960), or sodium citrate (BD363083)] were also used. Serum tubes were maintained in an upright position at 4°C for 1-2 hr to allow for coagulation prior to centrifugation. For plasma separation, blood samples were processed within 2-6 hours after collection. Plasma and serum samples were centrifuged in a swinging bucket rotor at 1,260 xg for 100 minutes at room temperature with low acceleration and no brake. Plasma phase was aspirated carefully from the top. Sera and plasma were sub aliquoted into cryo-vials and stored at −80°C.

### VAMS Sample Collection, Storage and Extraction

Volumetric absorptive microsampling (VAMS) (Mitra Collection Kit; Neoteryx, CA) was performed following the manufacturer’s instructions. Prior to sample collection, the lateral portion of the participant’s finger was cleaned with an alcohol swab and punctured with a lancet device provided in the kit. A hydrophilic 30-μL VAMS microsampler was held against the blood drop until filled. Two microsamplers were utilized per subject. Blood-filled microsamplers were returned to the protective cartridges, which were placed in sealed containers with silica desiccant packets and stored at room temperature for up to 2 weeks from the collection date. One microsampler tip (30 μl) was added to 300 μl VAMS buffer (1x PBS) (Corning, Manassas, VA), supplemented with 1% bovine serum albumin (Roche Diagnostics, Mannheim, Germany) and 0.5% Tween 20 (Sigma, MO) in a 1 ml-deep 96-well plate (Greiner Bio-One, Monroe, NC). The plate was covered with an adhesive seal and maintained shaking at 250 rpm for 16 hours at 4°C. The resulting eluates were heat-inactivated at 56°C for 60 minutes and clarified by centrifugation at 3,500 rpm for 5 minutes. Supernatants were used for ELISA or aliquoted and stored at -80°C.

## RESULTS

### The SARS-CoV-2 S1 RBD antigen used in this study

The assays described in this report were performed using a novel gp70-fusion protein form of the S1 RBD antigen. The gp70 domain possesses chaperone-like qualities, and this fusion protein system has been shown to facilitate the correct folding and glycosylation of conformational subdomains of the HIV-1 gp120 glycoproteins and to efficiently express epitopes recognized by HIV-1 patient sera that are dependent on native structures (22-24). The structure and properties of the gp70-RBD antigen are described in **Fig. 2**. The gp70 carrier domain has a His8 affinity tag inserted near its N-terminus to facilitate purification and an HRV-3C protease cleavage signal (LEVLFQGP with a GS linker) inserted before the RBD sequence (aa 316-544 of the Wuhan sequence) to allow cleavage and removal of the carrier domain, if desired (**Fig. 2A**). The purity of the intact fusion protein and the isolated RBD domain is shown in **Fig. 2B** (the reference RBD antigen contains a His6 affinity tag and thus appears slightly larger than the cleaved product; lanes 3 and 4).

**Figure 2.**
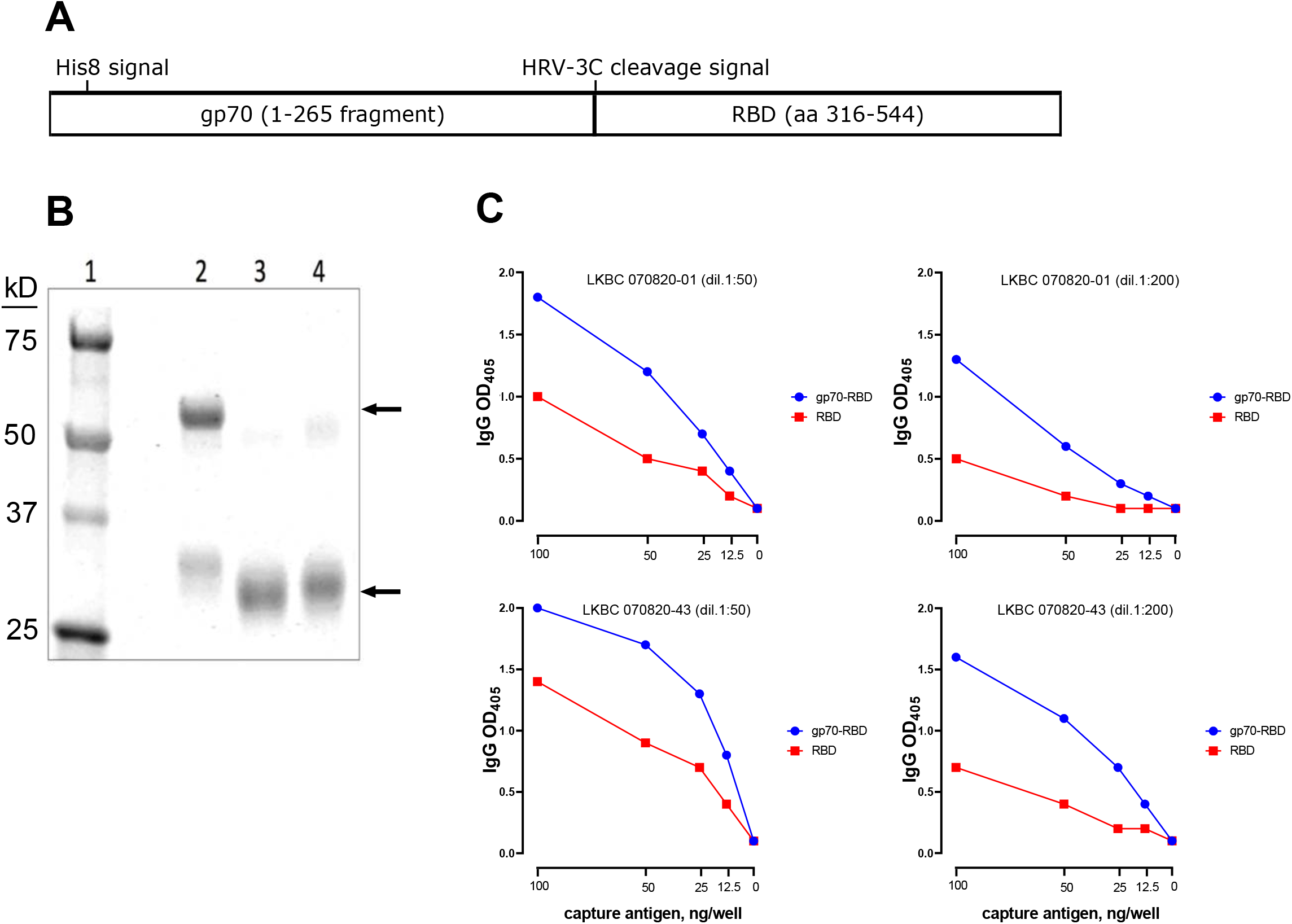
Design and activity of S1 RBD fusion protein. **(A)** Design of the gp70-RBD fusion protein. **(B)** SDS-PAGE analysis. Lane 1: Molecular weight markers; lane 2: gp70-RBD fusion protein; lane 3: RBD domain isolated from the fusion protein after proteolytic removal of the gp70 carrier domain by cleavage with HRV 3C protease; lane 4: reference RBD antigen from BEI resources. The relevant bands in each lane are marked by arrows. **(C)** Binding curves comparing equal amounts of the gp70-RBD fusion protein (blue symbols) and reference RBD antigen (red symbols) for reactivity against two convalescent sera used at two dilutions (1:50, left panels; 1:200, right panels) dil, dilution.

When we compared binding curves obtained utlizing equal protein concentrations of gp70-RBD fusion protein and reference RBD antigen against two convalescent sera (**Fig. 2C**), we observed that, despite its larger molecular weight, the fusion protein yielded a more sensitive signal than the reference RBD protein with both sera, under all antigen concentrations and serum dilutions tested. This result is likely due to more efficient binding of the antigen to the ELISA plate wells and better exposure of RBD epitopes resulting from the presence of the gp70 tag.

### S1 RBD-based antibody binding assay

The limited sequence conservation between the SARS-CoV-2 S1 RBD with that produced by non-pathogenic human coronaviruses (7) is expected to minimize the potential detection of cross-reactive antibodies. Indeed, pre-COVID sera did not react with SARS-CoV-2 S1 RBD but reacted with a mixture of non-SARS-CoV-2 coronavirus N antigens, presumably due to exposure to non-pathogenic human coronaviruses (**Fig. 3A**). RBD-specific IgG antibodies were detected in sera from convalescent subjects who had previously tested positive to SARS-CoV-2 PCR (n=83), hospitalized COVID-19 patients (n=146), and subjects fully vaccinated with COVID-19 RNA vaccines (n=283) (**Fig. 3B**). The overall higher reactivity of the convalescent group relative to the hospitalized patients is presumably due to a larger proportion of recently infected subjects in the latter group who may not have seroconverted. As expected (20, 25), the antibody response to mRNA vaccination was generally stronger than that to natural infection. No antibodies were detected in the negative control subjects [pre-COVID-19 (n=104) and SARS-CoV-2 PCR-negative subjects (n=103) that remained uninfected for at least 16 weeks after the blood draw tested in the assay (21) (**Fig. 3B**). In addition, our assay provided an accurate estimate of exposure to SARS-CoV-2 in 148 residents of Lakewood, NJ during the first peak of the COVID-19 pandemic in New Jersey (March-June 2020). The Lakewood township experienced one of the highest COVID-19 burdens in the US (12,800 cases per 100,000 – based on NJ Department of Health data (nj.gov/health) on COVID-19 as of 5/30/2021, and 2019 census estimates). All cases of SARS-CoV-2 infection in Lakewood were self-reported at a time when access to COVID-19 PCR testing was limited. The presence of seronegative subjects in this group (high burden, **Fig. 3B**) is consistent with the expected limitations of self-reporting. Taken together, these data demonstrate that detection of anti-RBD antibodies is highly suitable for determining exposure and estimating the prevalence of SARS-CoV-2 infection.

**Figure 3.**
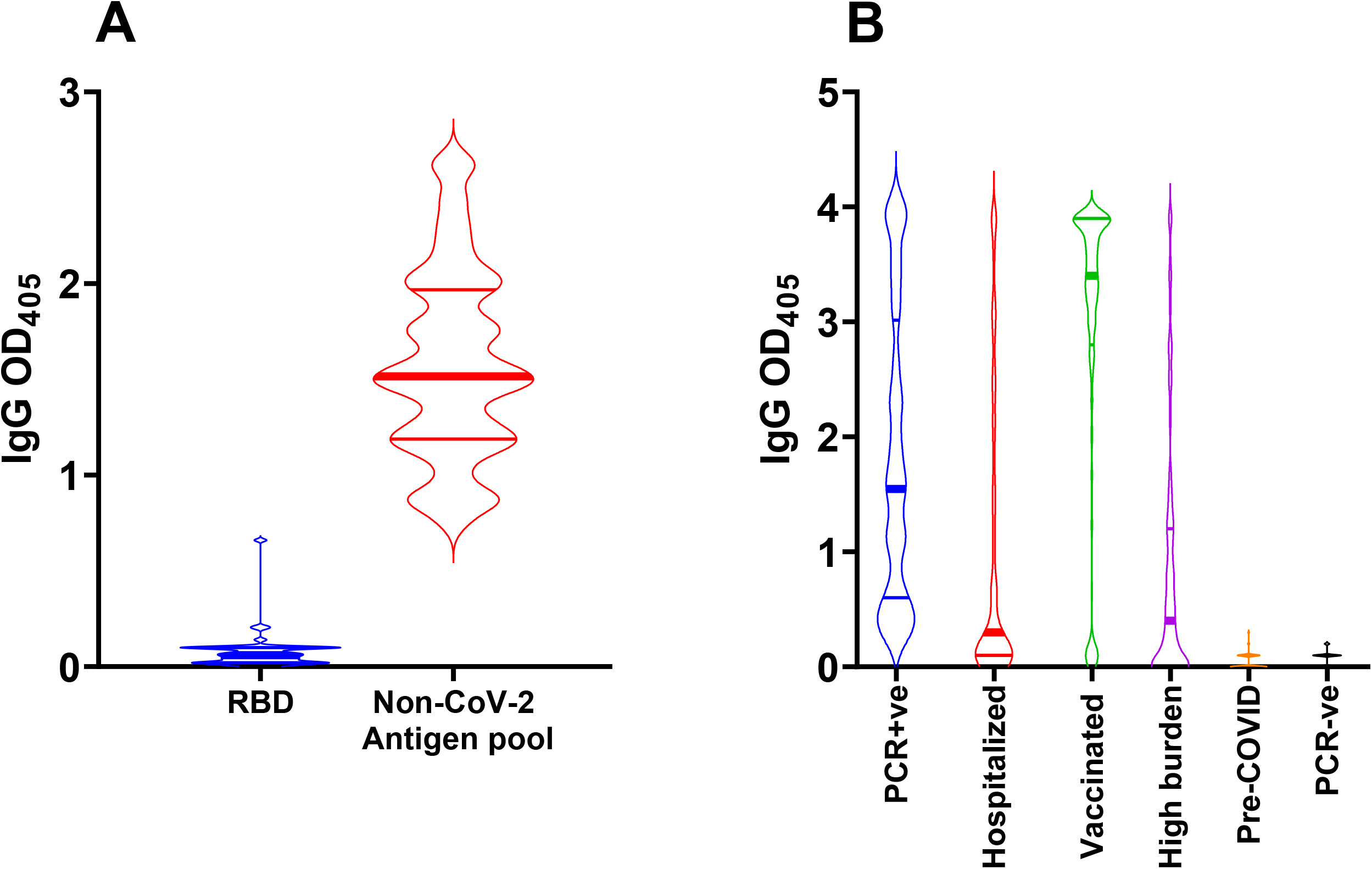
S1 RBD antibody binding assay. **(A)** Violin plots showing IgG reactivity of pre-COVID-19 sera (n=19) to RBD (blue) and a pool of non-SARS-CoV-2 coronavirus antigens (red) determined by ELISA using sera diluted 1:20. **(B)** Violin plots showing the reactivity to RBD of serum/plasma samples (diluted 1:80) from convalescent subjects who had tested PCR-positive for SARS-CoV-2 infection (n = 83, blue); COVID-19 hospitalized subjects (n=146, red); subjects who received full COVID-19 vaccination (n = 283, green); adult residents in the Lakewood, NJ township (high burden) s(n = 148, purple); pre-COVID-19 samples (n = 104; orange); SARS-CoV-2 PCR-negative subjects (n = 103; black). In all panels, the solid horizontal lines represent the median (thick line) and interquartile range (thin lines).

### Comparison with commercial serological assays for diagnosis of SARS-CoV-2 infection

We compared detection of virus-specific antibody responses to SARS-CoV-2 infection by our RBD-based ELISA vis-à-vis two assays (Roche Elecsys Anti-SARS-CoV-2, Basel, Switzerland, and Abbott Architect SARS-CoV-2 IgG assay, Chicago, IL, USA) that use SARS-CoV-2 N antigen as the capture reagent. Both assays have been authorized for emergency use by the US Food and Drug Administration and widely utilized in clinical settings during the pandemic. For this comparison, we used samples obtained from donors having PCR-confirmed infection prior to the introduction of SARS-CoV-2 vaccines (n=30) and pre-COVID-19 samples (n=70). For the commercial assays, cut-off values for positive or negative test determination were as established by the manufacturers. For our ELISA, the cut-off was established as the mean OD_405_ + 3SD value obtained with independent samples from SARS-CoV-2 PCR-negative subjects (n=103) that remained SARS-CoV-2 PCR-negative for at least 16 weeks (21) (rightmost group in **Fig. 3B**). The results of the three parallel assays are shown in **Fig. 4**. While the sample size was somewhat limited, the results suggest that our RBD-based ELISA has excellent sensitivity and specificity (**Fig. 4**) and compares favorably with commercial assays that are widely utilized in clinical settings.

**Figure 4.**
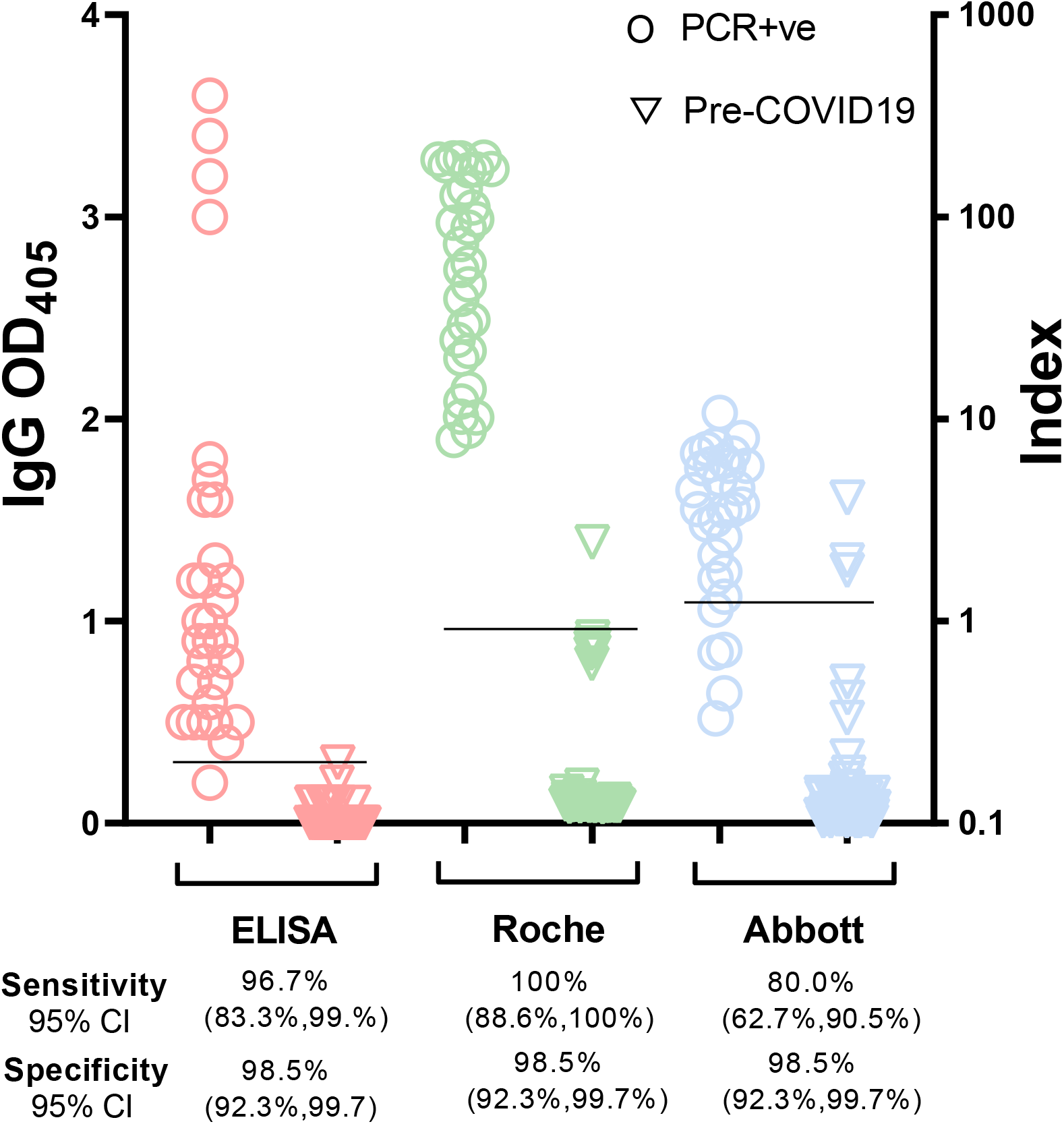
Comparison among three serological assays. The dot plots show IgG results obtained with our in-house automated ELISA (RBD-based) (pink symbols), the Roche Elecsys^®^ Anti-SARS-CoV-2 assay (N-based) (green symbols), and the Abbott Architect SARS-CoV-2 IgG assay (N-based) (blue symbols). Serum/plasma samples used for the three-way comparison were obtained from subjects who tested PCR-positive for SARS-CoV-2 infection (n = 30, open circles); pre-COVID-19 samples (n = 70, open triangles). Each symbol represents one study subject. Black horizontal lines represent cut-off values each serological assay. Cut-off values for the commercial assays were as per manufacturer’s instructions. For the in-house ELISA, the cut-off value (OD_405_ = 0.3) was calculated as the mean + 3 SD obtained with sera from 103 SARS-CoV-2 PCR-negative subjects who remained negative for at least 16 weeks after the blood draw utilized in the assay (data shown in Fig. 3B). It is noted that, for clarity purposes, a single scale (Index on the right y axis) was used for both commercial assays. However, the index calculation is different in the two assays; therefore, the relative numbers cannot be compared across assays. CI, confidence interval.

### Dried blood microsampling vs. phlebotomy for blood collection

Phlebotomy requires specialized personnel and specialized means of transporting blood tubes. However, studies conducted outside of healthcare settings, such as seroprevalence studies, require blood draws by non-specialized personnel or even by the study subjects themselves, possibly at remote sites. One such procedure involves microsampling by Mitra cartridges (https://www.neoteryx.com), which allows for collection of 10-50 ul of blood by finger stick, maintenance of the dried blood sample at room temperature for weeks (26) and, as needed, sample shipping to the testing site by regular mail. When we tested known SARS-CoV-2 seropositive (n=50) and seronegative subjects (n=12), we observed a clear separation between the two groups (**Fig. 5A**). In addition, when we tested in parallel blood samples collected using Mitra cartridges and phlebotomy from the same SARS-CoV-2 seropositive (n=13) and seronegative (n=3) subjects, we observed a strong correlation between the results obtained with blood samples drawn by the two methods (R^2^= 0.92 by Pearson correlation coefficient) (**Fig. 5B**). Thus, microsampling and phlebotomy can be used interchangeably for peripheral blood collection.

**Figure 5.**
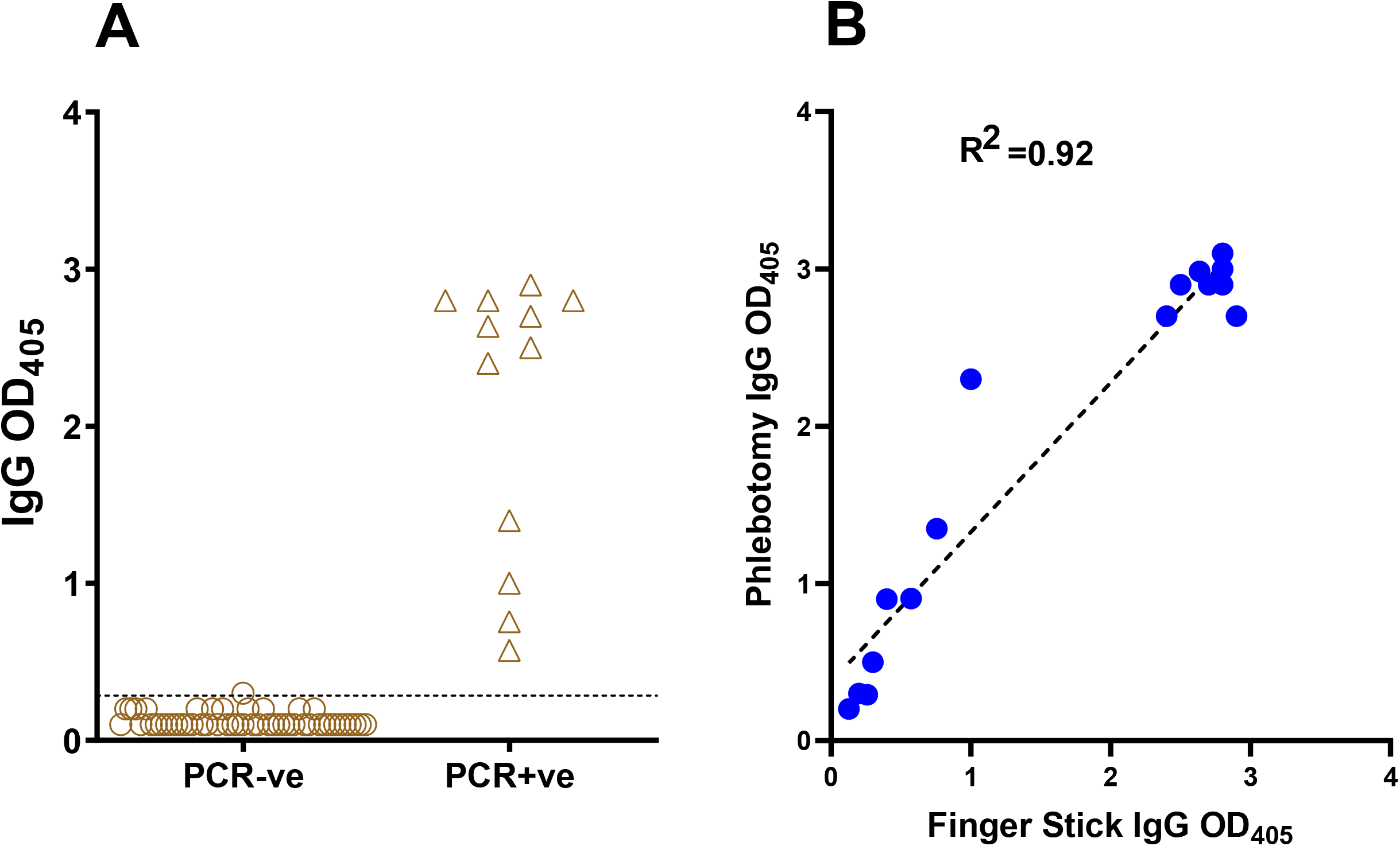
Comparison of dried blood microsampling and phlebotomy for blood collection. **(A) Dried-blood microsampling.** The dot plot shows RBD-specific IgG antibody binding determined by ELISA with samples collected by finger stick utilizing Mitra microsamplers from subjects that tested PCR-negative (n = 50, circles) or PCR-positive for SARS-CoV-2 infection (n = 12, triangles). The horizontal line represents the cut-off value of the assay, calculated as in the legend to Fig. 4. **(B) Correlation between ELISA results obtained from the same subjects by phlebotomy and by finger stick and Mitra microsampling**. The correlation plot shows results obtained with samples from 16 subjects (13 seropositive and 3 seronegative for SARS-CoV-2 infection). Pearson correlation coefficient (R^2^) of the comparison is also shown. In both panels, each symbol represents one study subject.

### Matrix equivalency assays

Since different study designs can result in collection of either serum or plasma from peripheral blood, we conducted a matrix equivalency test for our assay. Moreover, since plasma can be collected from blood collection tubes containing different anticoagulants, we also tested for equivalency of plasma obtained from different tubes. To perform these comparisons, we sampled in parallel serum obtained from a serum-separator tube (containing inert clot activator) and plasma obtained from blood collection tubes containing various types of anticoagulants (potassium EDTA, lithium heparin, sodium heparin, and sodium citrate) from five subjects that were negative for SARS-CoV-2 infection both by PCR and antibody assays. We used all serum and plasma matrices obtained from these negative subjects, and ELISA blocking buffer as a comparator, to dilute convalescent serum from a SARS-CoV-2-infected subject (PCR-positive and seropositive) to low (1:20), medium (1:80), and high (1:320) dilution. When we tested the resulting samples for anti-RBD antibody binding, we observed essentially no difference in ELISA values for each of the seropositive sample dilutions, regardless of the matrix used for dilution (**Fig. 6A**). Thus, utilization of serum or plasma from different blood collection tubes had no detectable effect on antibody binding results.

**Figure 6.**
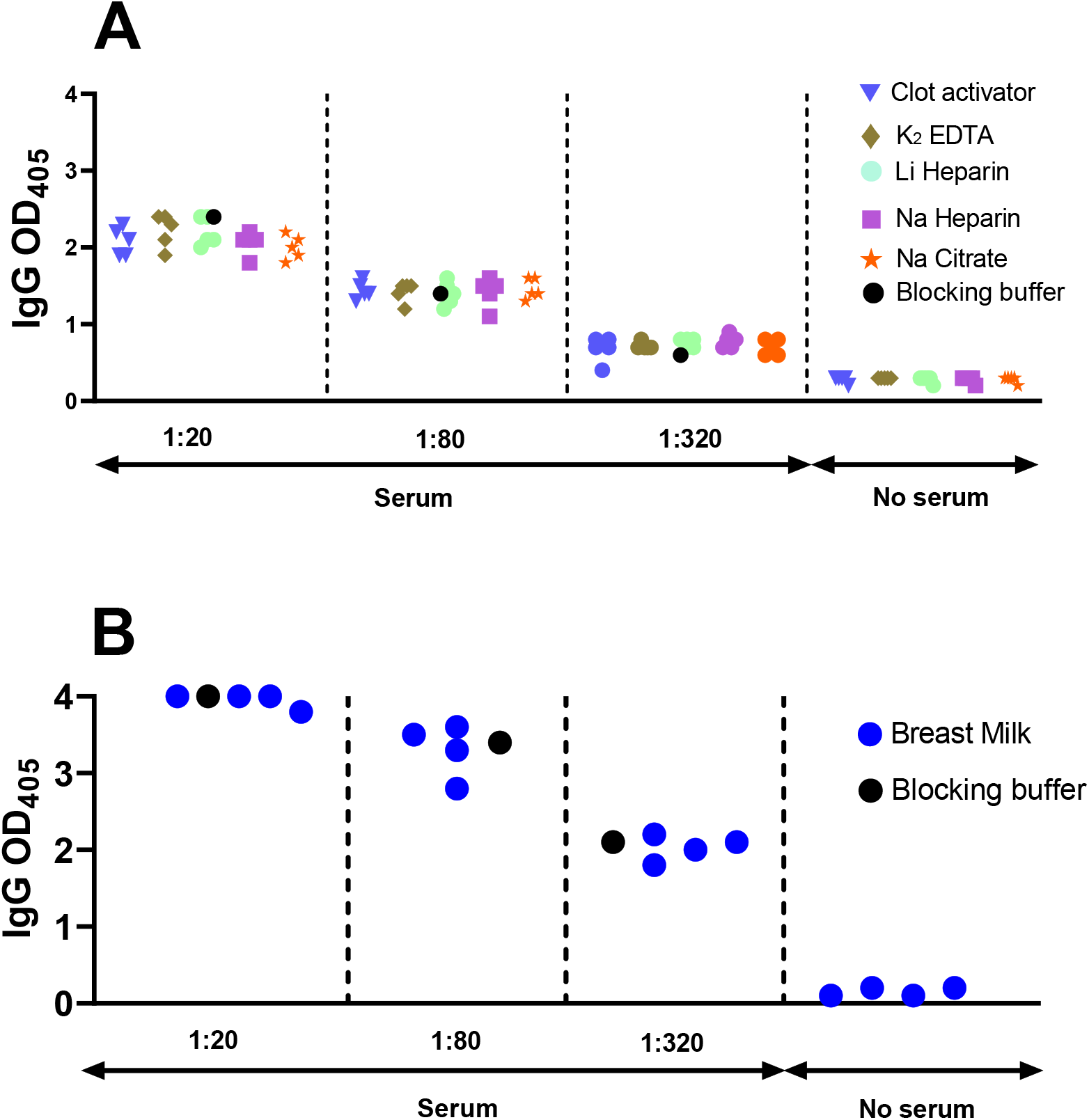
Matrix equivalency assays. The dot plot shows RBD-specific IgG antibody binding determined by ELISA. **(A) Serum and plasma matrices**. Serum and plasma samples were obtained from SARS-CoV-2 PCR-negative subjects (n = 5) using concurrently five different collection tubes, as indicated by the different symbols. **(B) Breast milk matrix**. Breast milk samples were collected from four SARS-CoV-2 PCR-negative lactating mothers. **(A**,**B)** All matrices (serum and plasma in panel A and breast milk in panel B) were used to serially dilute serum from one SARS-CoV-2 antibody positive subject at 1:20, 1:80, and 1:320, as indicated. The same seropositive sample was also diluted in conventional ELISA blocking buffer (reference condition, black circle). No serum indicates ELISA results obtained with the various matrices only (serum, plasma, breast milk), in the absence of the diluted seropositive serum. In both panels, each symbol represents one matrix per study subject.

Antibodies can be passively transferred from mother to baby through lactation. Regarding COVID-19, anti-SARS-CoV-2 antibodies have been detected in breast milk of mothers infected with SARS-CoV-2 or vaccinated against COVID-19, for example (27). We tested whether breast milk affects antibody detection by diluting convalescent serum from a SARS-CoV-2-infected subject in breast milk from four SARS-CoV-2 negative women, as described above for the plasma vs serum equivalency assay. We found that the ELISA readings obtained for breast-milk-diluted samples were almost identical with those obtained with the same sample, conventionally diluted in 2% non-fat milk (**Fig. 6B**). Collectively, the results indicate that our ELISA protocol is compatible with various matrices, including serum, plasma obtained using different anticoagulants, and non-blood bodily fluids such as breast milk.

## CONCLUSIONS

Monitoring the burden and spread of infection during the COVID-19 pandemic is of paramount importance, whether in small communities or large geographical settings. Serology, which detects the host antibody response to the infection, is the most appropriate tool for this task, since virus-derived markers are not reliably detected outside of the acute phase of the infection. Here we show that our S1 RBD-based ELISA is well suited to detect the antibody response to SARS-CoV-2 infection and to COVID-19 vaccination. We also provide a proof-of-principle demonstration of the value of COVID-19 seroepidemiology, since our assay can identify individuals who were infected with SARS-CoV-2 among those exposed or perceived to have been exposed because they live in a high-burden area. Thus, notwithstanding immune impairment limiting the sensitivity of the immunoassay, seroepidemiology has the potential to yield more accurate estimates of prevalence of infection than epidemiological tools based on clinical symptomatology or reported exposure.

We also demonstrate that our ELISA is accurate, versatile, and highly suited for research and clinical applications. Our protocol is performed utilizing robotic sample handling and dilution and automated ELISA. Moreover, it compares favorably with accurate commercial tests that have been widely used in clinical practice to determine exposure to SARS-CoV-2. Furthermore, our protocol accommodates use of various blood- and non-blood-derived biospecimens as well as dried blood obtained with microsampling cartridges that are appropriate for remote sampling and transportation.

Additionally, the suitability for samples obtained in small volumes constitutes a distinct advantage of our in-house, fully automated ELISA over commercial assays widely used in clinical settings, which require relatively large volumes (typically ≥100 μl per single assay) to accommodate the dead volume of the system. Thus, our RBD-based ELISA protocols are uniquely suited for seroepidemiology and other large-scale studies requiring parsimonious sample collection outside of healthcare settings.

## Data Availability

All raw data will be provided on request.

## Acknowledgements

We thank the PHRI biosafety officers and the RBHS Institutional Biosafety committee for fast-track review and approval of laboratory protocols and practices related to handling of SARS-CoV-2 and infected biospecimens; the Rutgers Institutional Review Board for timely review and approval of COVID-19-related protocols for human subject protection; Daniel Fine and Steven Libutti for supporting the start of our COVID-19 work; Nancy Reilly and the entire Rutgers Corona Cohort team that established a cohort from which some of the serum samples utilized in this study were obtained; and Dennis Burton and Pei-Yong Shi for providing biological reagents. This work was funded by NIH grants R01 HL149450, R01 HL149450-S1, U01 AI122285-S1, P30 ES005022, K23 AR070286, and UL1 TR003017.

